# Leveraging regulatory monitoring data for quantitative microbial risk assessment of *Legionella pneumophila* in cooling towers

**DOI:** 10.1101/2024.05.19.24307585

**Authors:** Émile Sylvestre, Dominique Charron, Xavier Lefebvre, Emilie Bedard, Michèle Prévost

## Abstract

Cooling towers are critical engineered water systems for air conditioning and refrigeration but can create favorable conditions for *Legionella pneumophila* growth and aerosolization. Human exposure to *L. pneumophila*-contaminated aerosols can cause Legionnaire’s disease. Routine monitoring of *L. pneumophila* in cooling towers offers possibilities to develop quantitative microbial risk assessment (QMRA) to guide system design, operation, control, and maintenance. Here, we used the regulatory monitoring database from Quebec, Canada, to develop statistical models for predicting *L. pneumophila* concentration variability in cooling towers and integrate these models into a screening-level QMRA to predict human health risks. Analysis of 105,463 monthly *L. pneumophila* test results revealed that the exceedance rate of the 10^4^ colony forming unit (CFU) per liter threshold was constant at 10% from 2016 to 2020, emphasizing the need to better validate the efficacy of corrective measures following the threshold exceedances. Among 2,852 cooling towers, 51.2% reported no detections, 38.5% had up to nine positives, and 10.2% over ten. The gamma or the lognormal distributions adequately described site-specific variations in *L. pneumophila* concentrations, but parametric uncertainty was very high for the lognormal distribution. We showed that rigorous model comparison is essential to predict peak concentrations accurately. Using QMRA, we found that, to meet a health-based target of 10^-6^ DALY/pers.-year for clinical severity infections, an average *L. pneumophila* concentration below 1.4 × 10^4^ CFU L^-1^ should be maintained in cooling towers. We identified 137 cooling towers at risk of exceeding this limit, primarily due to the observation or prediction of rare peak concentrations above 10^5^ CFU L^-1^. Effective mitigation of those peaks is critical to controlling public health risks associated with *L. pneumophila*.

## 1 Introduction

Exposure to *Legionella pneumophila* through inhalation of aerosols produced by engineered water systems, such as cooling towers, is a common cause of Legionnaire’s disease. This disease can result in a severe form of pneumonia, particularly threatening individuals who are older, have weakened immune systems, or suffer from chronic lung diseases (National Academies of Sciences and Medicine, 2020).

Cooling towers, engineered water systems designed to remove excess heat from buildings by cooling water, are critical for air conditioning and refrigeration but can provide ideal conditions for the growth and aerosolization of *L. pneumophila*. These systems operate using evaporative cooling. Warm water from cooling systems — usually between 29 and 35 °C, a temperature range favorable for *L. pneumophila* growth — is sprayed as fine droplets onto packing or honeycomb material. As ambient air is drawn into the cooling tower, either by natural or mechanical ventilation, a small portion of the water evaporates, reducing its temperature but also generating aerosol droplets that may carry *L. pneumophila*. While cooling towers should be equipped with drift eliminators designed to minimize aerosol droplets released into the atmosphere, the efficiency of these devices is not absolute (ASHRAE, 2008). The release of contaminated droplets into the atmosphere can pose public health risks for neighboring communities.

The growth of *L. pneumophila* colonies in cooling towers is influenced by various factors, such as the presence of protozoa (which serve as hosts), biofilms, nutrient availability, water temperature, and manufacturing materials (Kusnetsov et al., 1993; Paniagua et al., 2020; Türetgen and Cotuk, 2007). To minimize growth, the primary strategy involves chemical water disinfection (Kim et al., 2002). However, predicting the presence and survival of *L. pneumophila* in cooling towers solely from design and operational parameters is challenging. Various risk management guidelines and guidance documents based on the hazard analysis and critical control point (HACCP) method recommend routine monitoring of *L. pneumophila* in bulk water to validate water treatment efficiency (ASHRAE, 2018; 2020; Cooling Technology Institute (CTI), 2000; World Health Organization (WHO), 2011). Legal requirements for routine monitoring of *L. pneumophila* have been established across numerous countries (Radziminski and White, 2023; Van Kenhove et al., 2019), often necessitating corrective actions when specified *L. spp or L. pneumophila* concentrations are exceeded. For example, Quebec’s regulation requires intervention and confirmation of the effectiveness of corrective measures when the *L. pneumophila* concentration exceeds 10^4^ colonies forming unit (CFU) L^-1^ (Gouvernement du Québec, 2014).

A quantitative microbial risk assessment (QMRA) framework has been proposed to assess health risks associated with exposure to *L. pneumophila*-laden aerosols from cooling towers (Hamilton et al., 2018). This framework incorporates dose–response models to predict human health effects from the deposition of *L. pneumophila* in the lungs (Armstrong and Haas, 2007) and uses a Gaussian plume model to simulate the fate and transport of contaminated aerosol droplets (Hardy et al., 2006). Applying QMRA makes it possible to estimate public health risks based on concentrations of *L. pneumophila* in the bulk water from cooling towers. These risk predictions can be compared with health-based targets to provide a foundation for risk-based criteria to guide system design, operation, control, and maintenance (National Academies of Sciences and Medicine, 2020).

Within the QMRA framework proposed by Hamilton et al. (2018), the health risk is directly proportional to the concentrations of *L. pneumophila* in the bulk water of the cooling tower. Extensive routine *L. pneumophila* monitoring data from programs designed to validate treatment efficiency offer a significant opportunity to develop statistical models to investigate temporal variations in these concentrations. Parametric models, such as mixed Poisson distributions, have been widely used to model temporal variations in microbial concentrations in surface water sources (Haas et al., 1999; Masago et al., 2004; Teunis et al., 1997). These distributions have also been used in ecology to describe variations in the abundance of populations governed by an environmental carrying capacity (Dennis and Patil, 1988; Dennis and Patil, 1984). Despite the apparent potential of such models, their application to model routine monitoring *L. pneumophila* data remains unexplored. Bridging this gap could facilitate the development of more transparent, risk-based strategies for *L. pneumophila* risk assessment and management.

The objectives of our study are to i) develop candidate statistical models to predict the variability and uncertainty in *L. pneumophila* concentrations obtained from routine monitoring of bulk water in cooling towers, ii) establish a framework for model comparison and selection and implement it for an extensive database, and iii) incorporate selected models within a screening-level QMRA model to predict human health risks associated with exposure to droplet aerosols generated by a representative cooling tower.

## 2 Methodology

### 2.1 Database

In Quebec, *L. pneumophila* monitoring is required by the regulation for the maintenance of cooling towers (Gouvernement du Québec, 2014). We obtained the Quebec sregulatory database, which includes *L. pneumophila* monitoring results for 2852 cooling towers (1960 buildings) in Quebec, Canada. For each cooling tower, the database consists of results from monthly monitoring of *L. pneumophila* concentrations in bulk water obtained yearly or on a seasonal basis from 2016 to 2020. It also includes general system information, including the building type, usage type, service period, and location. Specific information on water treatment (types of biocides used, application frequencies, dosages, etc.) was unavailable. The regulation defines an action level of 10^4^ colony-forming units (CFU) L^-1^ and a human health risk level of 10^6^ CFU L^-1^. Upon exceedance of the action level, the regulation requires immediate corrective intervention. A strict decontamination procedure must be applied when the concentration of *L. pneumophila* is equal to or greater than 10^6^ CFU L^-1^. Additional *L. pneumophila* results following a decontamination procedure were included in the database for some cooling towers, following testing requirements. However, these results could not be differentiated from routine monitoring results.

### 2.2 Sample collection and *L. pneumophila* enumeration

Each sample was collected and stored following Standard DR-09-11 (Centre d’expertise en analyse environnementale du Québec, 2022). *L. pneumophila* enumeration was performed using culture-based methods adapted from AFNOR NF T90-431 or ISO 11731:2017, depending on the laboratory conducting the analyses. Diluted samples were spread on glycine vancomycin polymyxin cycloheximide agar medium (GVPC) or BMPA, with or without acid (pH 2; 5 minutes) or heat (50 °C; 30 minutes) pre-treatment or a combination of acid and heat. Samples were then incubated at 36 °C for 8-11 days, and presumptive colonies of *Legionella* spp. were grown with buffered charcoal yeast extract (BCYE) supplemented or not with cysteine at 36 ± 1.5 °C for 3-5 days. *L. pneumophila* species were identified, in most cases, by a latex agglutination test, and all results were expressed as CFU. Results at the detection limit were reported in CFU L^-1^ calculated as 1 CFU per volume of sample tested.

### 2.3 Estimation of *L. pneumophila* counts from reported concentrations

Statistical inference was not made directly from reported concentrations since treating detection limits as actual microbial concentrations or as censored data can significantly bias statistical analyses (Chik et al., 2018).

We developed an approximation method to estimate the colony numbers and assayed water volumes for each reported concentration. Initially, for concentrations reported at the detection limit, we calculated the tested volume (*V*_*i*_) for each instance (*i*) using the formula:

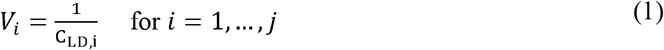

where C_LD,i_ represents the detection limit and *j* is the count of these instances in the database. Subsequently, for each detected concentration (C_detected,m_), we computed the corresponding number of colonies (*k*_*i,m*_) by applying the previously determined volume (*V*_*i*_) in the equation:

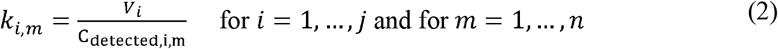

Here,*n* is the total number of distinct detected concentrations. This calculation resulted in *j* pairs of colony numbers and volumes, each associated with a specific detected concentration.

To approach the statistical analysis conservatively and avoid underestimating the sampling uncertainty, we selected the minimum integer of colonies (*k*_*min*_) along with its corresponding volume (V). This approach minimizes the colony count to ensure the broadest possible confidence intervals for the average number of colonies per sample.

### 2.4 Estimation of *L. pneumophila* concentration from counts

A discrete random variable can characterize the distribution of organisms within a specific volume of water. Assuming that these organisms are randomly distributed in the water sample, the probability of finding a specific count of organisms (*k*) within a sample of known concentration (*c*) and volume (*V*) is given by the Poisson distribution:

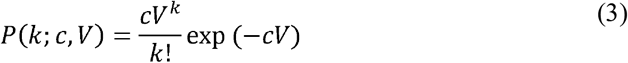

For a sample adhering to a Poisson distribution, the maximum likelihood estimate for the concentration (*c*) is the sample arithmetic mean given by:

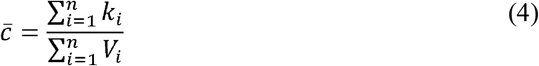

This estimator is only valid if organisms are distributed “randomly,” i.e., the bulk solution is well mixed, and organisms are not aggregated.

### 2.5 Temporal stability of the *L. pneumophila* concentration

When a series of samples is collected at a regular interval, obtained results may indicate that the concentration is stable or variable over time. To assess whether the concentration of *L. pneumophila* is stable over time in the cooling tower, we used a likelihood ratio test to evaluate the fit of a Poisson distribution to observed data (Haas et al., 1999). The null hypothesis of the test posits that the data set is adequately described by a single Poisson distribution with a constant concentration,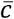. The likelihood of the null hypothesis is given by:

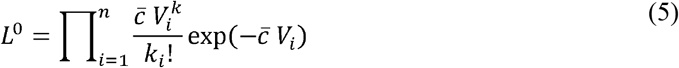

Conversely, the alternative hypothesis suggests that each sample has its concentration 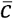 at the time of sampling. Therefore, the likelihood for the alternative hypothesis is:

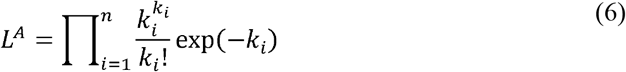

The test statistic (∧) can be simplified to:

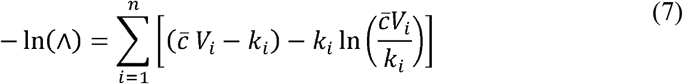

The null hypothesis is rejected when the value of −2In(∧) exceed the upper 1 − α percentile of a χ^2^ distribution with n − 1 degrees of freedom. The degrees of freedom represent the difference in the number of parameters between the alternative hand null hypotheses. The error risk α was set at 5%.

### 2.6 Temporal variability in *L. pneumophila* concentrations

Distributions of microorganisms in water are often more dispersed than what a Poisson distribution would predict; that is, the variance of the number of organisms exceeds the mean. This overdispersion relative to the Poisson distribution can result from spatial or temporal heterogeneities. This heterogeneity can be accounted for by a continuous random variable representing the concentration of each sample. The marginal distribution of the number of organisms is then obtained by the following integral:

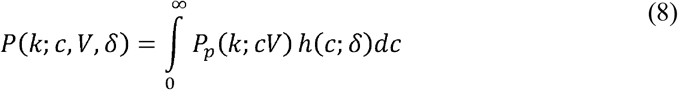

where *h* is a continuous distribution with parameters δ representing the temporal variations in concentrations. Gamma and lognormal distributions have been selected to model temporal variations. The general properties of these distributions are presented in Table 1.

**Table 1.** Density functions describing the variation of a concentration c, mean, and standard deviation for the gamma and lognormal distributions.

| Distribution | Density function | Arithmetic mean | Standard deviation |
| --- | --- | --- | --- |
| Gamma ($\alpha, \beta$) | $\frac{\beta^\alpha}{\Gamma(\alpha)} c^{\alpha-1} e^{-\beta c}$ | $\alpha/\beta$ | $\alpha/\beta^2$ |
| Log-normale ($\mu, \sigma$) | $\frac{1}{c\sigma\sqrt{2\pi}} \exp\left[-\frac{[\ln c - \mu]^2}{2\sigma^2}\right]$ | $\exp\left(\mu + \frac{\sigma^2}{2}\right)$ | $\sqrt{\exp(\sigma^2) - 1} \exp\left(\mu + \frac{\sigma^2}{2}\right)$ |

### 2.7 Bayesian inference

Parameters for Poisson and mixed Poisson distributions were inferred using Bayesian models. The hierarchical model structure is outlined as follows: At the first level, the observed number of colonies per volume is distributed according to a Poisson distribution. At the second level, concentration *c* is modeled as a latent variable (i.e., not directly observable) following a continuous distribution (either gamma or log-normal for this study), making the first level conditional on *c*. The hierarchical structures for Poisson-gamma and Poisson-log-normal models are:

- Poisson gamma: *x*_*i*_ | *c*_*i*_, *V*_*i*_ ∼ Poisson (*c*_*i*_ *V*_*i*_), *c*_*i*_ ∼ Gamma (*α, β*)
- Poisson log-normal: *x*_*i*_ | *c*_*i*_, *V*_*i*_ ∼ Poisson (*c*_*i*_ *V*_*i*_), *c*_*i*_ ∼ Lognormal(*μ, σ*)

Prior distributions were chosen to ensure Markov chain stationarity while minimizing the prior influence on the posterior distribution. A conjugate Gamma(*α, β*) prior was assigned to parameter *c*, with α and β set to 0.01. Uniform (min, max) priors were used to infer gamma distribution parameters α and β, with bounds set at 0 and 10 for α, and 10^-12^ and 10^-1^ for β. A uniform (min, max) prior was selected for μ, with bounds at − 10 and 10. An (λ) prior was allocated to σ as suggested by McElreath (2018), with λ set at 0.1, assuming the logarithm of the standard deviation was significantly below 50 for all cooling towers.

Models were fitted using Markov chain Monte Carlo (MCMC) simulations with rjags (v4-10) in R (v4.1.0). For each parameter, three Markov chains were run for 10^5^ iterations following a burn-in of 10^3^ iterations. The Brooks-Gelman-Rubin scale reduction factor was applied to assess chain convergence, and the effective sample size (the ratio of sample size to autocorrelation in Markov chains) was evaluated to ensure comprehensive exploration of the posterior distribution, deemed well-estimated at an effective size over 10,000. Brooks-Gelman-Rubin reduction factors and effective sample sizes were calculated using the diagMCMC function (Kruschke, 2014).

### 2.8 Model comparison and selection

The fit quality of Poisson and mixed Poisson distributions was compared using the deviance information criterion (DIC) (Spiegelhalter et al., 2002). The DIC is computed as follows:

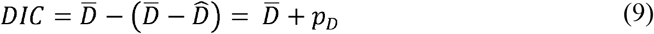

where 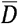 is the mean deviance across the sampled parameter values from the posterior distribution, and 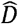 is a penalty term related to the risk of model overfitting. A model with a lower DIC value is considered superior. Generally, a model with a DIC at least three points lower than that of other models is significantly better (Spiegelhalter et al., 2002). For hierarchical models with a latent variable (e.g., mixed Poisson distributions), different DICs can be calculated for the model’s hierarchical levels. The fit of a Poisson distribution should be assessed with a conditional DIC (cDIC), while that of a mixed Poisson distribution should be evaluated with a marginal DIC (mDIC) (Millar, 2009). The cDIC can be directly calculated using the current version of rjags in R. However, computing the mDIC is more complex as it requires integrating the likelihood function of the distribution, which cannot be done directly in rjags. Thus, mDICs for mixed Poisson distributions were computed through numerical integration following the approach proposed by Quintero and Lesaffre (2018).

The cooling towers were classified using a decision-making algorithm identifying the best model describing *L. pneumophila* concentrations (Fig. 1). For cooling towers where the statistical distribution of data could be validated, we computed the median value and the 95% uncertainty interval of the average *L. pneumophila* concentration predicted by the best-fit model. The analysis was conducted in R (version 4.3.0). The R code used for data analysis and visualization can be found in the GitHub repository [NOTE: URL will be provided in the final paper].

**Fig 1.**
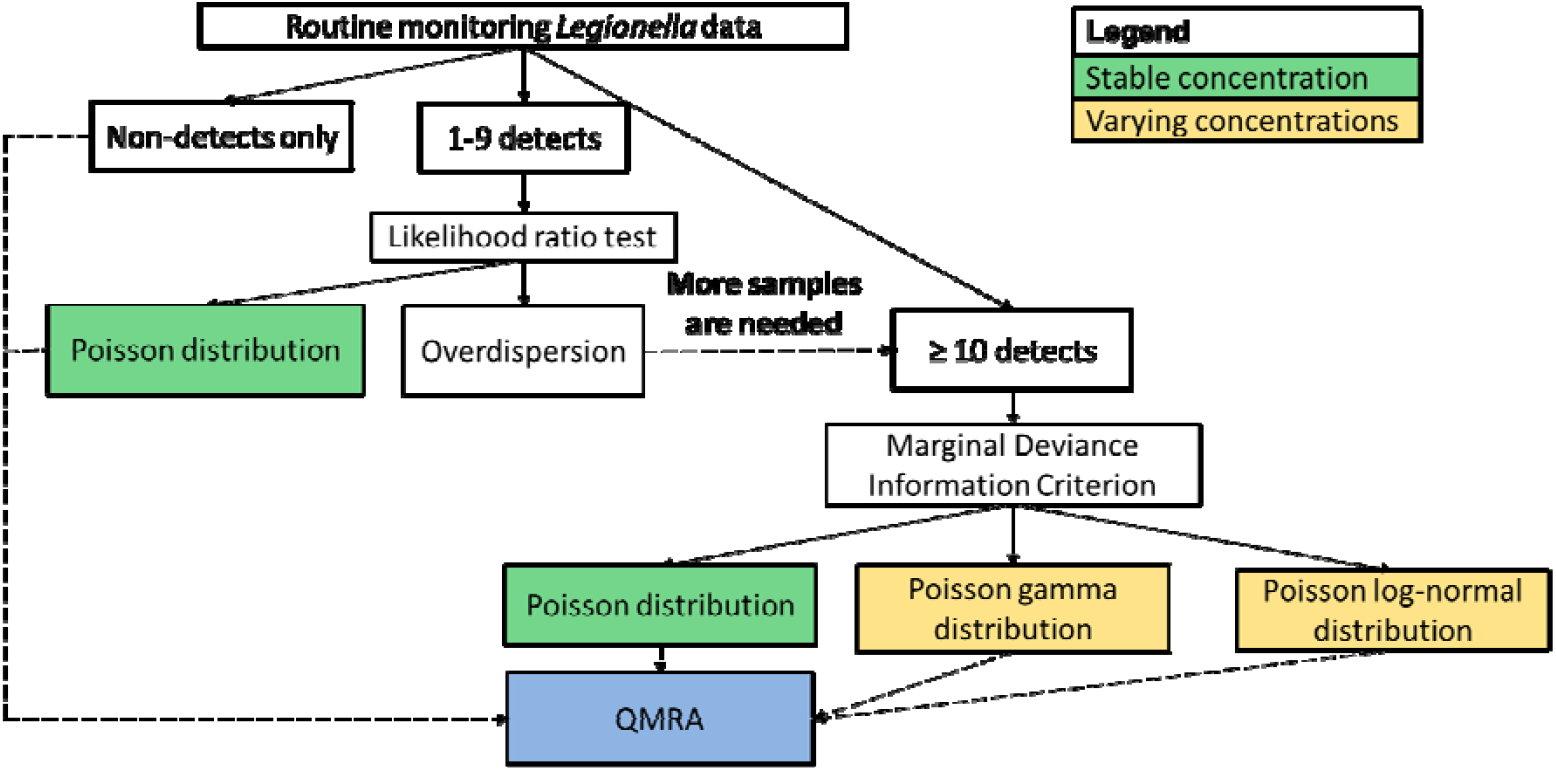
Decision algorithm to determine the best fit model to assess *L. pneumophila* concentrations in bulk water from a cooling tower.

### 2.9 Screening-level quantitative microbial risk assessment

To determine the dose of *L. pneumophila* that can deposit in the respiratory tract of the exposed subject, it is necessary to develop and integrate a series of sub-models that account for various stages of the process. These stages include the emission of contaminated droplets by the cooling tower, the transport and atmospheric dispersion of the droplets, the inactivation of *L*.

*pneumophila* during atmospheric transport, and its inhalation and deposition in the human respiratory tract (Table S1). For our screening-level QMRA, a hypothetical cooling tower was considered, characterized by an effective head of 10 m and an average recirculation flow rate of 10^3^ L s^-1^. The analysis also assumes that a drift eliminator reduces evaporative water loss to 0.003% of the recirculation flow rate (ASHRAE, 2008).

#### 2.9.1 Emission, fate, and transport of aerosol droplets discharged by the cooling tower

We focused our analysis only on aerosol droplets with diameters ≤ 150 μm, as they quickly evaporate once airborne and can be transported over long distances. To estimate the mass-weighted fraction of these aerosols, we applied the lognormal distribution of aerosol mass modelled by Peterson and Lighthart (1977) from measurements obtained at the cooling tower outlet by Shofner and Thomas (1971). For the specific fractions of *L. pneumophila* bacteria transferred from water to aerosols with a diameter of 1-10μm, we relied on estimates from Hamilton et al. (2018) extracted from the empirical cumulative distribution function obtained by Allegra et al. (2016). These fractions are conservative for QMRA, given that the enumeration of *L. pneumophila* bacteria for each droplet diameter was determined using a qPCR method, which measures both viable and dead cells.

We employed a Gaussian model to model the transport and dispersion of these aerosols. Based on results obtained at 66 meteorological stations in Quebec by Ilinca et al. (2003), we considered an arithmetic mean annual wind speed of 4.5 m s^-1^. For environmental conditions, the Gaussian model predicts that the highest concentration of the pollutant emitted at an effective height of 10 m would be observed at a distance of 110 m from the point of emission.

Droplets were considered as evaporated immediately upon emission from the cooling tower. To evaluate the survival of *L. pneumophila* within these evaporated aerosol droplets, we used a first-order two-phase inactivation model developed from the data of Katz and Hammel (1987). This model predicts a high inactivation rate in the first 30 seconds (λ_El_= 0.12 s^-1^), which then decreases (λ_E2_=7×10^-4^ s^-1^). Inactivation rates for aqueous aerosols are expected to be very low (λ_A_ ∼ 1×10^-4^ s^-1^) compared to the rates for evaporated aerosols (first phase) (Hamilton et al., 2018).

#### 2.9.2 Inhalation, deposition of aerosols, and health effects

We used deposition efficiencies for aerosols with diameters ranging from 1 to10 μm obtained by Heyder et al. (1986) for an oral inhalation rate of 15 L air min^-1^, a respiratory cycle duration of 8 s, and an average respiratory volume of 1 L. Our model assumed an exposure duration of one hour per day at a frequency of 365 days per year, as proposed by Hamilton et al. (2018) to assess the risks associated with residential exposure. The dose – response models developed by Armstrong and Haas (2007) based on animal data from Muller et al. (1983) and Fitzgeorge et al. (1983) were used to predict infection with subclinical severity and infection with clinical severity. Clinical severity was defined as an infection requiring medical attention or seeking health services. The health impact of a clinically severe infection was assessed using the disability-adjusted life years (DALY) factor calculated by van Lier et al. (2016) based on surveillance data for Legionnaires’ disease in the Netherlands (Dijkstra et al., 2010; Dijkstra et al., 2008).

#### 2.9.3 Risk characterization

The health outcome target corresponds to the tolerable risk associated with exposure to *L. pneumophila* in contaminated aerosols a cooling tower produces. Infection risks and DALYs specific to each cooling tower in the database were assessed based on the arithmetic mean concentration of *L. pneumophila* in the water, estimated by parametric modeling. Risk estimates were compared to health-based targets of 1) 10^-4^ infections per person per year (Macler and Regli, 1993) and 2) 10^-6^ DALYs per person per year (Havelaar and Melse, 2003; World Health Organization (WHO), 2008).

## 3 Results

The database aggregated 105,463 monitoring results documenting *L. pneumophila* concentrations in 2,852 cooling towers from 1,960 buildings in Quebec, Canada. Empirical complementary cumulative distribution functions (CCDFs) of monitoring results grouped by year offer insights into exceedances of *L. pneumophila* concentration thresholds (Fig. 2). The 10^4^ CFU L^-1^ threshold has a steady exceedance probability of 10% over the 2016-2020 period. The exceedance probability of the 10^6^ CFU L^-1^ threshold also remains constant at around 0.5% for the whole period. Notably, the 10^7^ CFU L^-1^ threshold is exceeded more often in 2018 than in the other years.

**Fig 2.**
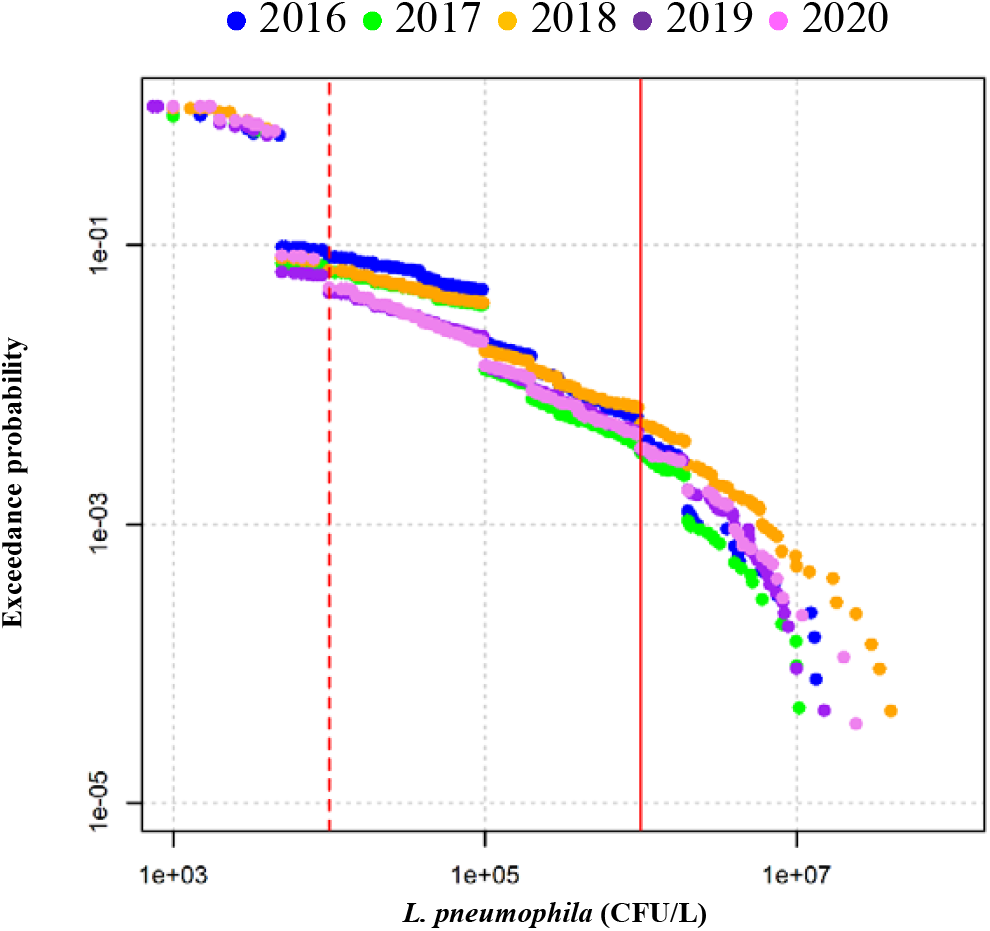
Empirical complementary cumulative distribution function of *L. pneumophila* concentrations in bulk water from 2852 cooling towers in Quebec, Canada, from 2016 to 2020. The dotted and solid lines indicate concentrations of 10^4^ and 10^6^ CFU L^-1^, respectively.

The 5-year arithmetic mean concentration and the maximum concentration for each cooling tower were computed to assess their relationship. The log-log relationship between the maximum and the arithmetic mean concentrations demonstrates that peak concentrations can substantially impact the 5-year arithmetic mean (Fig. 3). The arithmetic mean is typically about 1.0 log lower than the sample maximum.

**Fig 3.**
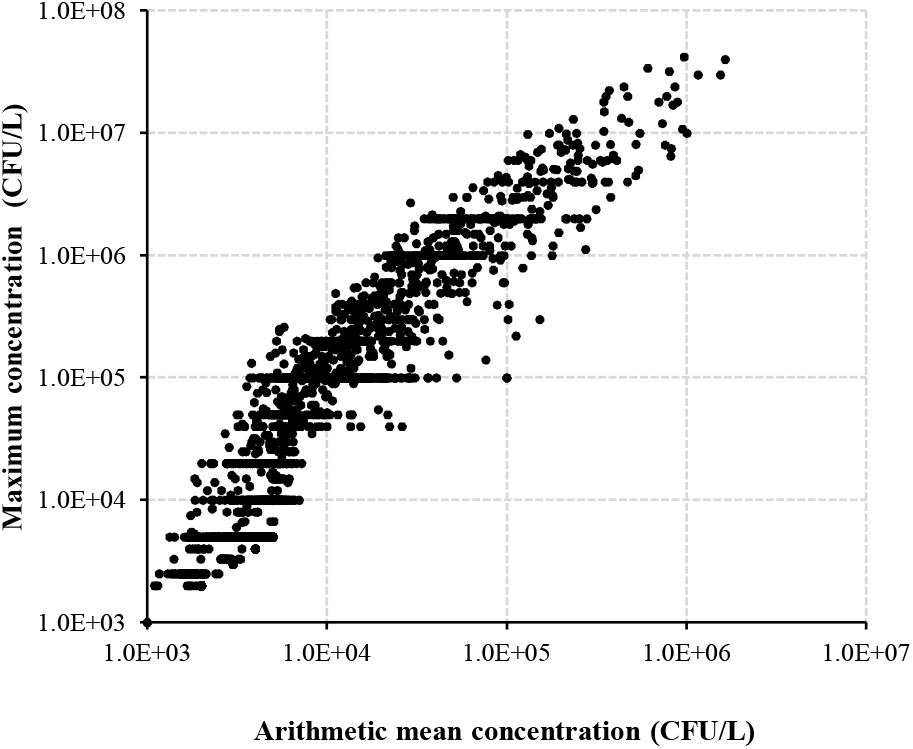
Relationship between the sample arithmetic mean concentration and the maximum concentration of *L. pneumophila* in bulk water from 2852 cooling towers in Quebec, Canada.

For 1461 cooling towers (about half of the total), all monitoring results are non-detects (Table 2). For 1,099 cooling towers, 1 to 9 positive results are obtained. Results from the Poisson test reveal that *L. pneumophila* concentrations are statistically stable for roughly 50% of these cooling towers, equivalent to around 20% of the total. For the remaining 50%, the limited number of positive results hinders accurate parameter estimation of the Poisson Gamma and Poisson lognormal distributions. Temporal variations could be assessed for the 292 cooling towers where the positive results count is ten or more (about 10% of the total). Within this subset, the marginal deviance information criterion (mDIC) suggests the Poisson distribution as the most suitable model for ten cooling towers, indicating stable concentrations despite consistent positive findings. For the remaining 282 cooling towers, the mDIC can discriminate between the Poisson gamma and Poisson lognormal for 145 cooling towers, often favoring the Poisson lognormal. For the remainder, the mDIC indicates a similar quality of fits.

**Table 2.** Classification of cooling towers based on the decision algorithm to determine the best-fit model to assess *L. pneumophila* concentrations in bulk water, as shown in Fig. 1. Models are the Poisson distribution (Pois.), the Poisson gamma distribution (PGA), and the Poisson lognormal distribution (PLN).

| Model | No positives | 1-9 positives | | $\geq 10$ positives | | | | Total |
| --- | --- | --- | --- | --- | --- | --- | --- | --- |
|  | n/a | Pois. | No pois. | Pois. | PGA | PLN | Mixed Poisson<sup>A</sup> |  |
| Nb of cooling towers | 1461 | 552 | 547 | 10 | 55 | 90 | 137 | 2852 |
| Fraction of total | 51.2% | 19.3% | 19.1% | 0.3% | 1.9% | 3.1% | 4.8% | 100% |

<sup>A</sup> Discrimination between the fit of the PGA and PLN is not possible because the difference in DIC between the two models is less than three points.

When results are Poisson distributed, arithmetic mean concentrations are consistently below 10^4^ CFU L^-1^ (Table 3). Predictions from the Poisson gamma generally indicate arithmetic mean concentrations below 10^5^ CFU L^-1^. Arithmetic mean concentrations surpass 10^6^ CFU L^-1^ in 30 of the 90 cooling towers with optimal Poisson lognormal fits. This discrepancy underscores the significant impact of distribution selection on average concentration estimates. Moreover, the span of the 95% uncertainty interval of the arithmetic mean predicted by the Poisson lognormal generally extends 1.0-to 2.0-log more than that of the Poisson gamma (Table S2, Figure 4). The CCDFs show that the predictions of the lognormal distribution can extrapolate beyond the maximum observation, unlike the gamma distribution. The uncertainty interval of the CCDF remains stable for the gamma distribution but not for the lognormal distribution, expanding as exceedance probabilities diminish. For certain cooling towers, like Cooling tower D, the CCDF exhibits distinct tail behaviors at exceedance probabilities below 1% (Figure 4). In such cases, the limited data size fails to adequately capture the upper tail’s behavior, explaining the often-similar mDICs for Poisson gamma and Poisson lognormal fits.

**Table 3.** Classification of 844 cooling towers based on their arithmetic mean *L. pneumophila* concentration in bulk water. The average concentration was predicted using the best-fit model determined using the decision algorithm shown in Fig. 1. The models are the Poisson distribution, the Poisson gamma distribution (PGA), and the Poisson lognormal distribution (PLN).

| Model | Arithmetic mean *L. pneumophila* concentration (CFU/L) |  |  |  | Total |
| --- | --- | --- | --- | --- | --- |
| | $< 10^4$ | $10^4 - 10^5$ | $10^5 - 10^6$ | $> 10^6$ | |
| Poisson (1-9 positives) | 552 (100%) | 0 (0.0%) | 0 (0.0%) | 0 (0.0%) | 552 (100%) |
| Poisson ($\geq 10$ positives) | 10 (100%) | 0 (0.0%) | 0 (0.0%) | 0 (0.0%) | 10 (100%) |
| PGA | 11 (20.0%) | 35 (63.6%) | 9 (16.3%) | 0 (0.0%) | 55 (100%) |
| PLN | 15 (16.6%) | 26 (28.8%) | 19 (21.1%) | 30 (33.3%) | 90 (100%) |
| Mixed Poisson<sup>A</sup> - PGA | 81 (59.1%) | 48 (35.0%) | 8 (5.8%) | 0 (0.0%) | 137 (100%) |
| Mixed Poisson<sup>A</sup> - PLN | 45 (32.8%) | 34 (24.8%) | 26 (18.9%) | 32 (23.3%) | 137 (100%) |

<sup>A</sup> Discrimination between the fit of the PGA and PLN is not possible because the difference in DIC between the two models is less than three points.

**Fig 4.**
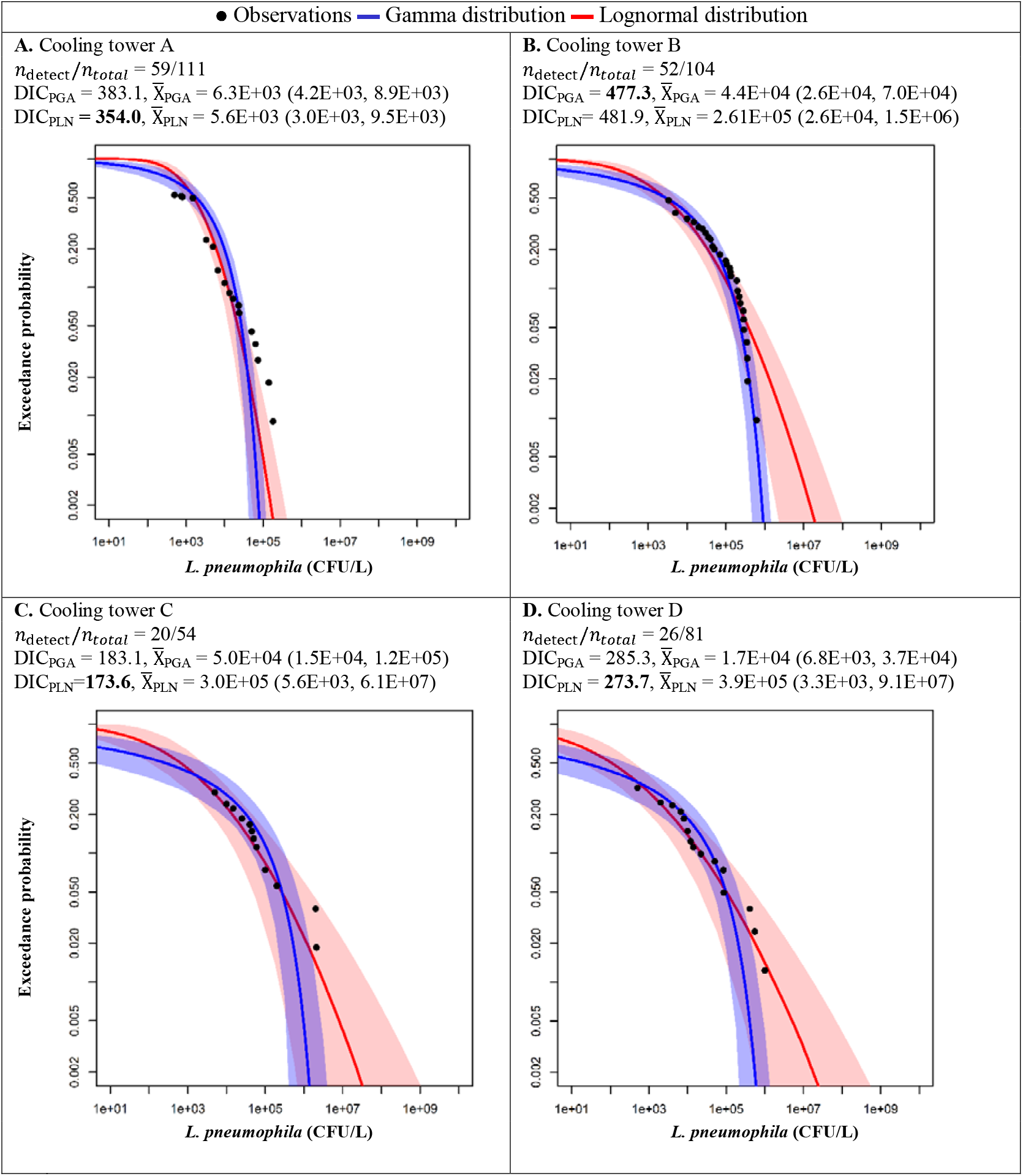
Complementary cumulative distribution function of the Poisson-gamma (PGA) and Poisson lognormal (PLN) distributions fitted to *L. pneumophila* data from four cooling towers from the Quebec database. The dark blue and red lines represent the best fits of the gamma and lognormal distributions, respectively. The blue and red areas represent the 95% uncertainty intervals of the gamma and lognormal distributions, respectively. The values of the marginal deviance information criterion (mDIC) and the arithmetic mean concentration (with 95% uncertainty interval) in the bulk water of the cooling tower (CFU/L) predicted by each model are listed. mDICs in bold indicate the best-fit models.

QMRA results indicate that achieving a health-based target of 10^-4^ inf./person-year requires maintaining the arithmetic mean *L. pneumophila* concentrations below 1.0E+03 CFU L^-1^ for the subclinical infection severity model and below 1.4E+06 CFU L^-1^ for the clinical infection severity model (Table 4). The stricter health-based target of 10^-6^ DALY/pers.-year results in a critical arithmetic mean concentration of 1.4E+04 CFU L^-1^. Results from the screening-level QMRA of 844 cooling towers are shown in Table 5. For both the arithmetic mean risk and its 97.5% uncertainty bound, this table documents the number of cooling towers that achieve the target or not. For cooling towers where the discrimination between the Poisson gamma and Poisson lognormal was not possible, risks were estimated for both models. Approximately 77% of those 844 cooling towers meet the 10^-6^ DALYs/pers.-year target. Using the 97.5% uncertainty bound as an input for QMRA lowers compliance to about 70%. Among the 562 cooling towers with stable concentration (Poisson distributed), the arithmetic mean risk and its upper uncertainty bound are generally below targets. For the Poisson gamma, roughly half of the cooling towers meet the 10^-6^ DALYs/pers.-year. The remaining cooling towers exhibit arithmetic mean risks about 1.0-log above this target. Risks predicted by the Poisson lognormal tend to surpass 10^-6^ DALYs/pers.-year, with 60 cooling towers with risks exceeding the target by more than 2.0-log and 137 cooling towers with the uncertainty bound exceeding the target by more than 2.0-log.

**Table 4.**
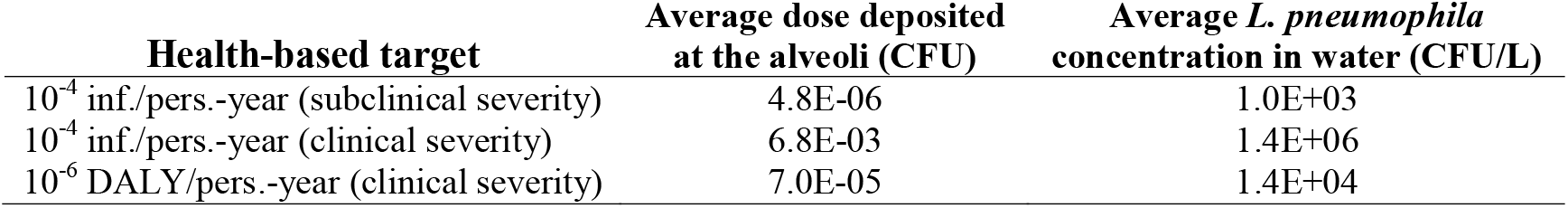
Critical arithmetic mean *L. pneumophila* dose deposited at the alveoli and critical arithmetic mean *L. pneumophila* concentration in water of the cooling water to achieve three different annual health-based targets. Parameter values of the QMRA model used to calculate the doses and concentrations are presented in Table S1.

**Table 5.**
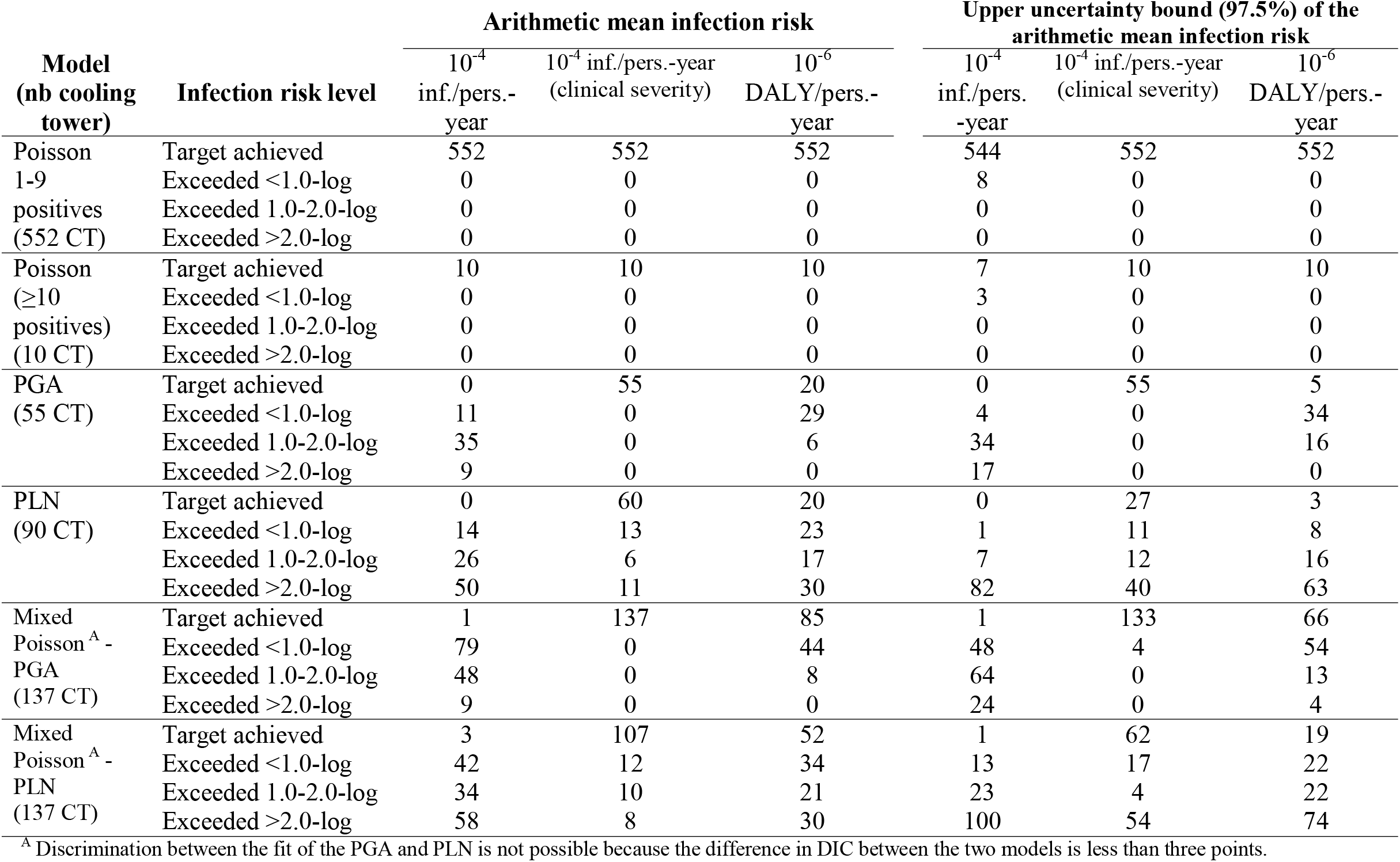
Classification of 844 cooling towers (CTs) based on their arithmetic mean infection risk and their upper uncertainty bound (97.5%) of the arithmetic mean infection risk. Risks have been calculated using a screening-level quantitative microbial risk assessment (QMRA). Risks are compared to three different annual health-based targets. Models are the Poisson distribution, the Poisson gamma distribution (PGA), and the Poisson lognormal distribution (PLN).

## 4 Discussion

### 4.1 Evolution of *L. pneumophila* concentration exceedance rates in Quebec

In Quebec, the period from July 2014 to June 2017 saw a documented decrease in exceedance rates of 10^4^ CFU L^-1^ for approximately 300 cooling towers (Racine et al., 2019), coinciding with the introduction of the new regulation (Gouvernement du Québec, 2014). The average annual exceedance rate decreased from 15% in 2014-2015 to 9% in 2016-2017. For the 2016-2020 period, our results indicate similar exceedance rates of about 10% for concentrations of 10^4^ CFU L^-1^ for the combined results of the 2852 cooling towers included in the database. The exceedance rates for concentrations of 10^5^ CFU L^-1^ and 10^6^ CFU L^-1^ also remained stable during this period, except for a notable increase in the exceedance rate of 10^7^ CFU L^-1^ in 2018. This increase may have been influenced by the extreme heat waves of summer 2018, potentially leading to increased cooling tower usage and operating conditions favorable to *L. pneumophila* growth. That summer was recorded as the hottest in 146 years of meteorological observations in southern Quebec (Lebel et al., 2019). Analyzing risk factors, such as air temperature, humidity level, and precipitation, could provide insights into the conditions that resulted in these extreme concentrations. Overall, the introduction of Quebec’s regulation appears to have initially reduced the high concentrations of *L. pneumophila* in Quebec’s cooling towers but did not reduce the exceedance rates of 10^4^ CFU L^-1^ to the lower values of around 1% found in the Canadian federal cooling tower databases (data not shown). Gathering specific information on the water treatment strategies used in these cooling towers, including the types of biocides, application frequencies, and dosages, could shed light on the causes of the differences observed between these datasets.

### 4.2 Variations in *L. pneumophila* concentrations in cooling towers over five years

To assess temporal variations in *L. pneumophila* concentrations in cooling towers, three discrete parametric distributions — Poisson, Poisson gamma, and Poisson lognormal — were proposed. We chose these discrete distributions to avoid replacing non-detects with a specific concentration (e.g., one organism per analyzed volume). Instead, we treated the sample concentration as a random variable that can be estimated using a Poisson distribution. This approach requires knowledge of the number of CFUs and the water volume analyzed per sample. However, laboratories do not typically report this critical information. We introduced a method to estimate the number of CFUs and the analyzed volume of each sample from reported concentrations, although the potential bias in these estimates could not be assessed. A direct comparison between the fit of these distributions to actual and estimated results would provide insights into the limitations of our approach. Ideally, laboratories should report raw data (i.e., the number of CFUs and the analyzed volume) to facilitate modelling of temporal variations for risk assessment.

We developed an algorithm to identify the best modelling approach based on monitoring results. Using a goodness-of-fit test for the Poisson distribution, we determined whether the concentration was statistically stable or variable in cooling towers when at least one positive result was recorded. The distribution of microorganisms in water typically exhibits more dispersion than the Poisson distribution predicts, meaning that the variance of organism counts exceeds their average number. Our analysis attributed overdispersion relative to the Poisson distribution solely to temporal variations in concentrations. Yet, other heterogeneity sources — such as organism aggregation in samples or variable recovery rates from enumeration methods — can also contribute to overdispersion (Haas and Heller, 1986). Although these other heterogeneity sources have not been analyzed for *L. pneumophila*, they are well-documented for other bacteria (El-Shaarawi et al., 1981; Haas and Heller, 1986; Pipes et al., 1977). Examining subsamples from individual water samples from cooling towers could elucidate the impact of *L. pneumophila* aggregation on concentration variation.

Gamma et lognormal distributions were chosen as mixture distributions to characterize temporal variations in *L. pneumophila* concentrations. The theoretical foundation for selecting these distributions lies in their ability to approximate random variations in population size, constrained by an environmental carrying capacity. The gamma distribution can be derived from population growth following the logistic function (Dennis and Patil, 1984), while the lognormal distribution emerges from the Gompertz function (Dennis and Patil, 1988). Both logistic and Gompertz models yield sigmoid functions representing population growth in three stages: initial slow growth, followed by optimal growth, which eventually slows down as the population approaches its carrying capacity. However, the Gompertz curve tapers off more gradually than the logistic curve. Thus, the lognormal distribution may indicate that *L. pneumophila* has not reached its carrying capacity, whereas the gamma distribution suggests that the carrying capacity has been reached. This carrying capacity might be affected by operational factors, including water treatment practices and the efficacy of interventions following regulatory threshold exceedances. Our analysis reveals that neither the gamma nor the lognormal distribution consistently outperforms the other in predicting variations. Identifying the most accurate model was not possible for many cooling towers, likely due to limited sample sizes. When discrimination between the gamma and lognormal is not feasible, we recommend selecting the lognormal to conservatively predict peak concentrations.

### 4.3 Challenges in monitoring *L. pneumophila* concentrations in cooling towers

Using static parametric distributions for modelling *L. pneumophila* concentrations assumes the stochastic process is stationary (i.e., the process’s structure is constant over time) and ergodic (i.e., the sample size is sufficiently large to reflect the process’s structure). However, fulfilling these conditions in cooling towers presents challenges. A shift in the average concentration due to changes in water treatment may render the process non-stationary, potentially requiring different analyses for pre- and post-intervention data. Additionally, our findings reveal that monthly monitoring over five years may not adequately represent the process. This is evidenced by the considerable parametric uncertainty in the *L. pneumophila* concentration distribution of some cooling towers. This result highlights the importance of accounting for short-term variations to predict public health risks. The causes of these fluctuations, such as *L. pneumophila* growth events or biofilm detachment, remain uncertain, limiting precise recommendations for monitoring. Nevertheless, our analysis suggests that the monitoring guidelines and regulations require revision for the risk management of high-risk cooling towers (e.g., those exceeding the 10^4^ CFU L^-1^ threshold). A more frequent monitoring interval, potentially bi-weekly or weekly, notably during periods of higher risk like warmer months, could be key to managing public health risks associated with these cooling towers.

### 4.4 Advancing QMRA for systems with high variability

QMRA was initially developed to predict health risks associated with enteric pathogen concentrations in surface water. In such systems, the variability in concentrations is generally low enough to accurately estimate the long-term average pathogen concentration with low-frequency (e.g., monthly) monitoring over a set period (e.g., two years) (USEPA, 2010). Evaluating compliance with an annual health-based target can make sense for these systems. However, for systems with higher variability in pathogen concentrations, such as *L. pneumophila* in cooling towers, demonstrating compliance with an annual health-based target may not be feasible. For such systems, a short-term health-based risk target may be preferable, as previously recommended for drinking water safety management by Signor and Ashbolt (2009). Our reverse-QMRA model indicates the critical annual average concentration of *L. pneumophila* should be 1.4 × 10^4^ CFU L^-1^ to achieve a health-based target of 10^-6^ DALY/pers.-year, aligning closely with Quebec’s threshold of 10^4^ CFU L^-1^. Although the Quebec regulatory threshold was established “per sample,” our findings demonstrate that concentrations exceeding 10^4^ CFU L^-1^ can substantially increase the annual average concentration. Therefore, maintaining “per sample” concentrations below 10^4^ CFU L^-1^ could be an effective strategy for managing short-term clinical infection risks.

Adopting a short-term risk target could significantly impact how monitoring strategies are developed. For a “per sample” strategy to work, the monitoring frequency must be statistically valid, ensuring concentrations consistently remain below the set threshold with a defined confidence level. This approach demands detailed statistical analyses to determine the necessary monitoring frequency to capture *Legionella* growth, bloom, and sloughing events, as recommended by National Academies of Sciences and Medicine (2020). The validity of a risk-based threshold also depends on the assumptions underlying the QMRA model. Our study’s assumptions largely align with those of Hamilton et al. (2018). We diverged in assuming negligible reflection of viable bacteria from surfaces. Also, we found a fraction of aerosol mass with diameters of 100 μm or smaller of 17%, based on the distribution of Peterson and Lighthart (1977), which is above the 1.4% reported by Hamilton et al. (2018). Assessing aerosol mass distributions emitted by more recently constructed cooling towers could enhance the accuracy of exposure models.

### 4.5 Proactive strategies to reduce *L. pneumophila* exceedance thresholds

Under Quebec’s regulations, exceeding the 10^4^ CFU L^-1^ threshold requires identifying the causes of the increase, implementing corrective measures, and assessing the efficacy of these measures. However, the effectiveness of these interventions appears limited, given the recurring exceedances of the 10^4^ CFU L^-1^ and 10^6^ CFU L^-1^ thresholds since 2016. To address this issue, adopting high-frequency *L. pneumophila* monitoring following exceedances could enable early detection of peak concentrations, allowing for a better evaluation of the corrective measures. The implementation of comprehensive water management plans could shift focus from reactive to preventive measures, potentially reducing costs and resources associated with frequent monitoring following threshold exceedances.

## 5 Conclusions

The analysis of an regulatory database, including *L. pneumophila* concentrations monitored monthly from 2,852 cooling towers across Quebec, Canada, between 2016 and 2020, led to the following conclusions:

- The analysis of the 105,463 monitoring results shows that exceedance rates of the 10^4^ and the 10^6^ CFU L^-1^ thresholds have remained constant at 10% and 0.5%, respectively. While the implementation of Quebec regulations in 2014 initially reduced threshold exceedances, this trend was not sustained from 2016 to 2020. Establishing validation procedures for corrective actions to prevent recurring threshold exceedances could be an effective risk management strategy to address this issue.
- For the 2,852 cooling towers, 51.2% reported exclusively non-detects, 38.5% reported between one to nine positives, and 10.2% recorded more than ten positives. For this latter group, site-specific temporal variations in concentrations of *L. pneumophila* were often well described by either the gamma or the lognormal distribution. Due to the distinct behaviors of their upper tails, these distributions predicted considerably divergent arithmetic mean concentrations for some data sets. Implementing rigorous model comparison and selection approaches is essential to reliably predict peak concentrations.
- Our screening-level QMRA model suggests that maintaining a yearly average *L. pneumophila* concentration of 1.4 × 10^4^ CFU L^-1^ or lower is necessary to achieve a health-based target of 10^-6^ DALY/pers.-year for infections of clinical severity. QMRA identified 137 cooling towers at risk of exceeding this health-based target, primarily because of observed or predicted concentrations above 10^5^ CFU L^-1^. Therefore, maintaining “per sample” concentrations below 10^4^ CFU L^-1^ could be an effective strategy for managing short-term clinical infection risks. Increasing the monitoring frequency would be necessary to enhance the identification and mitigation of *L. pneumophila* growth periods in these cooling towers.

Detailed information on water treatment practices at cooling towers (e.g., biocide types, application frequencies, and dosages) would be valuable in investigating design and operational factors contributing to *L. pneumophila* proliferation within cooling towers.

## Data Availability

All data produced in the present work are contained in the manuscript.

## 6 Acknowledgments

This study was funded by the Natural Sciences and Engineering Research Council of Canada (NSERC) through the Industrial Chair on Drinking Water to M.P. Alliance Grant ALLRP 545363-19 to M.P., E.B. and Discovery grant RGPIN-2021-04341 to M.P.

The Alliance grant is funded by NSERC (66%) under the Alliance collaborative research partnership program and cofunded by Public Service and Procurement Canada, Régie du Batiment du Québec, Société Québécoise des Infrastructures, Centre d’expertise en analyse environnementale du Québec du ministère de l’Environnement, de la Lutte contre les changements climatiques, de la Faune et des Parcs, Bioalert solutions, and IDEXX.

We would like to thank Suzel Bourdeau from Régie du Batiment du Québec (RBQ), Carol-Ann Roy from Société Québécoise des Infrastructures (SQI), Jeff Moffat, and Clayton Truax from Public Services and Procurement Canada (PSPC) for providing us with these databases. We also thank Manuela Villion from Centre d’expertise en analyse environnementale du Québec du ministère de l’Environnement (CEAEQ) for assisting us in interpreting the data. Finally, we would like to thank Marwan Chacrone for his contribution to the analysis of the databases.

