## Supplementary material for "Leveraging regulatory monitoring data for quantitative microbial risk assessment of *Legionella pneumophila* in cooling towers"

**Table S1.** Input parameters of the quantitative microbial risk assessment model.

| **Equation** | **Parameter** | **Unit** | **Value** | **Reference** |
| --- | --- | --- | --- | --- |
|  | The concentration of the organism in water ($C_{water}$) | CFU/L | Specific | - |
| $Q_{emission}=\frac{1}{R}C_{water}Q_{water}E\times q$ | Detection method recovery rate ($R$) | fraction | 0.70 | Johnson et al. (2018)  McCuin et al. (2021) |
|  | Water recirculation rate ($Q_{water}$) | m^3^ /s | 10^3^ | Hensley (2009) |
|  | Effectiveness of drift eliminator ($E$) | % | 0.003 | ASHRAE (2008) |
|  | Mass-weighted fraction of aerosols with diameter ≤ 100 $\mu$m ($q$) | fraction | 0.17 | Peterson and Lighthart (1977)  Shofner and Thomas (1971) |
|  | Wind speed ($\mu)$ | m/s | 4.5 | Ilinca et al. (2003) |
| $C_{air}(x,y,z)=\frac{Q_{emission}}{2\pi\mu\sigma_{y}\sigma_{z}}\exp\left[ \left( -\frac{y^{2}}{2\sigma_{y}^{2}} \right) \right]\left\{ \exp\left[ -\frac{\left( z-H_{e} \right)^{2}}{2\sigma_{z}^{2}} \right]+\alpha\exp\left[ -\frac{\left( z+H_{e} \right)^{2}}{2\sigma_{z}^{2}} \right] \right\}\exp\left[ -\lambda\frac{x}{\mu} \right]$  where  $\sigma_{y}=R_{y}x^{r_{y}}$  $\sigma_{z}=R_{z}x^{r_{z}}$ | Stability parameter $R_{y}$ (class C) | - | 0.230 | Seinfeld (2020) |
|  | Stability parameter $r_{y}$(class C) | - | 0.855 | Seinfeld (2020) |
|  | Stability parameter $R_{z}$ (class C) | - | 0.076 | Seinfeld (2020) |
|  | Stability parameter $r_{z}$(class C) | - | 0.879 | Seinfeld (2020) |
|  | Center distance from emission source (x) | m | 110 | Assumed |
|  | Lateral distance from emission source (y) | m | 0 | Assumed |
|  | Height of inhalation zone (z$)$ | m | 1.5 | Assumed |
|  | Effective height of CT ($H_{e})$ | m | 10 | Assumed |
|  | Reflection term ($\alpha$) | - | -1 | Assumed |
|  | Inactivation rate of *L. pneumophila* in evaporated aerosols ($\lambda$) | s^-1^ | ≤ 30 s: 0.12  > 30 s: 7×10^-4^ | Katz and Hammel (1987) |

**Table S1 (cont’d)**. Input parameters of the quantitative microbial risk assessment model

| **Equation** | **Parameter** | **Unit** | **Value** | **Reference** |
| --- | --- | --- | --- | --- |
| $D=C_{air}It\sum_{i}^{n} {F_{i}DE}_{i}$ | Inhalation rate ($I$) | m^3^ minute^-1^ | 0.015 | Heyder et al. (1986)  USEPA (2011) |
|  | Duration of exposure ($t$) | Minute | 60 | Assumed |
|  | Fraction of organisms in aerosols of diameter $i$for $i$=1-10 $\mu$m ($F_{i}$)  1  2  3  4  5  6  7  8  9  10 | % | 17.50  16.39  15.56  6.67  3.89  2.50  2.78  5.00  5.28  3.89 | Allegra et al. (2016) |
|  | Deposition efficiency of diameter aerosols $i$ on the alveolus for $i$=1-10 $\mu$m (${DE}_{i}$)  1  2  3  4  5  6  7  8  9  10 | Fraction | 0.25  0.53  0.62  0.61  0.52  0.40  0.29  0.19  0.12  0.06 | Heyder et al. (1986) |
| (1) $P_{inf} =1-\exp\left( -rD \right)$  (2) $P_{\mathrm{DALY}} =\frac{\mathrm{DALY}}{\mathrm{infection}}P_{inf}$  (3) $P_{inf, an} =1-\left( 1-P_{inf} \right)^{N}$ | Dose-response parameter for *L. pneumophila*, probability of infection ($r_{inf}$) | - | 0.059 | Armstrong and Haas (2007)  Muller et al. (1983) |
|  | Dose-response parameter for *L.* pneumophila, infection with clinical severity ($r_{isc}$) | - | 4.1 × 10^-5^ | Armstrong and Haas (2007)  Fitzgeorge et al. (1983) |
|  | DALY for Legionnaires' disease (DALY/infection) | Fraction | 0.97 | van Lier et al. (2016) |
|  | Exposure frequency ($N$) | Day | 365 | Assumed |

**Table S2**. Classification of 844 cooling towers (CTs) based on the span of the 95% uncertainty interval of the arithmetic mean *L. pneumophila* concentration in bulk water. These results were obtained using the best-fit model determined using the decision algorithm shown in Fig. 1. The models are the Poisson distribution, the Poisson gamma distribution (PGA), and the Poisson lognormal distribution (PLN).

|  | **Span of the 95% uncertainty interval of the arithmetic mean *L. pneumophila* concentration** | | |  |
| --- | --- | --- | --- | --- |
| **Model** | < 1.0-log | 1.0–2.0-log | > 2.0-log | Nb of CTs |
| Poisson (1-9 positives) | 148 (26.8%) | 161 (29.2%) | 243 (44.0%) | 552 (100%) |
| Poisson (≥ 10 positives) | 10 (100.0%) | 0 (0.0%) | 0 (0.0%) | 10 (100%) |
| PGA | 38 (69.1%) | 17 (30.9%) | 0 (0.0%) | 55 (100%) |
| PLN | 5 (5.6%) | 10 (11.1%) | 75 (83.3%) | 90 (100%) |
| Mixed Poisson ^A^ - PGA | 95 (69.3%) | 42 (30.7%) | 0 (0.0%) | 137 (100%) |
| Mixed Poisson ^A^ - PLN | 13 (9.5%) | 22 (16.1%) | 102 (74.5%) | 137 (100%) |

^A^ Discrimination between the fit of the PGA and PLN is not possible because the difference in DIC between the two models is less than three points.
